# High prevalence of deep venous thrombosis in non-severe COVID-19 patients hospitalized for a neurovascular disease

**DOI:** 10.1101/2020.09.03.20187344

**Authors:** Olivier Rouyer, Irène Nora Pierre-Paul, Amadou Talibe Balde, Damaris Jupiter, Daniela Bindila, Bernard Geny, Valérie Wolff

## Abstract

**Introduction:** Severe SARS-CoV-2 infection, responsible for COVID-19, is accompanied by venous thromboembolic events particularly in intensive care unit. In non-severe COVID-19 patients affected by neurovascular diseases, the prevalence of deep venous thrombosis (DVT) is unknown. The aim of or study was to report data obtained after systematic Doppler ultrasound scanning (DUS) of lower limbs in such patients.

**Methods:** Between March 20 and May 2, 2020, consecutive patients with neurovascular diseases with non-severe COVID-19 were investigated with a systematic bedside DUS.

**Results:** Thirteen patients were enrolled including 10 acute ischemic strokes, one transient ischemic attack, one cerebral venous thrombosis and one haemorrhagic stroke. At admission the median National Institute of Health Stroke Scale (NIHSS) was of 6 (IQR, 0–20). We found a prevalence of 38.5% of asymptomatic calves’ DVT (n = 5) during the first week after admission despite thromboprophylaxis. Among them, one patient had a symptomatic pulmonary embolism. Two patients died during hospitalization but the outcome was favourable in the others with a discharge median NIHSS of 1 (IQR, 0–11).

**Discussion/Conclusion:** Despite thromboprophylaxis, systematic bedside DUS showed a high prevalence of 38.5% of DVT in non-severe COVID-19 patients with neurovascular diseases. Therefore, we suggest that this non-invasive investigation should be performed in all patients of this category.

## Introduction

The coronavirus disease 2019 (COVID-19) is characterized by a high prevalence of venous thromboembolism, particularly in patients with severe illness admitted in intensive care unit [1, 2]. Systematic venous lower limbs Doppler ultrasound scanning (DUS) in COVID patients showed a high prevalence of deep venous thrombosis (DVT) from 14.7% up to 85.4% [3, 4]. Furthermore, high pulmonary embolism rate in stroke patients with COVID-19 was recently reported [5], thus questioning the increased risk of thromboembolism in such cases. We aimed to determine the prevalence of DVT in non-severe COVID-19 patients hospitalized for a neurovascular disease using systematic bedside DUS of lower limbs in a stroke unit.

## Materials and Methods

Thirteen consecutive patients with neurovascular diseases and non-severe COVID-19, admitted in the stroke unit of Strasbourg University Hospital between March 20 and May 2, 2020, were investigated. COVID-19 was confirmed by RT-PCR and/or chest computed tomography (CT-scan) [6]. DUS of the entire lower limbs venous system was performed at admission and subsequently on days 7 and 14 by the same experimented practitioner (OR) using an IE33 Sonographer device (Philips, Bothell, WA, USA). The approval for this study was obtained from the local ethics committee of the Strasbourg University Hospital (reference CE-2020–111) and verbal informed consent was obtained from each patient. This study is recorded in http://clinicaltrials.gov. (Unique identifier: NCT04452422).

## Results

Patients’ characteristics are reported in the Table. Brain magnetic resonance imaging showed 10 ischemic strokes (76%), one cerebral venous thrombosis (8%), and was normal in one patient leading to the diagnosis of transient ischemic attack (8%). Brain CT-scan showed a cerebellar hematoma in one patient (8%). A large vessel occlusion was found in 5 patients and in 7 the carotid territory was affected. The remaining patient out of 13 had an occlusion of the ophthalmic artery. The patients received the following treatments: alteplase alone (n = 1), aspirin alone (n = 3), endovascular procedure alone (n = 2) or combined endovascular procedure with intra-arterial alteplase (n = 1) or intravenous alteplase (n = 3). Cerebral venous thrombosis was treated by effective anticoagulation, haemorrhagic stroke by reversal anticoagulation.

Nine patients received thromboprophylaxis and 3 received effective anticoagulation. One was not treated by anticoagulant because he could walk around.

The median National Institute of Health Stroke Scale (NIHSS) was of 6 (IQR, 0–20) at admission. The neurological outcome was favourable in 11 cases and the median NIHSS for discharge was of 1 (IQR, 0–11). One patient stayed five days in intensive care unit for monitoring because of a worsening dyspnoea. Two patients died because septicaemia complications for one and terminal phase of a newly discovered cancer.

DUS showed an acute asymptomatic calf DVT in 2 patients at admission and in 3 others at day 7. At day 14, no more thrombotic event was noticed. Among the 5 patients with DVT, one patient with a complicated pulmonary embolism was treated by effective anticoagulation, one patient continued the thromboprophylaxis treatment due to muscular calf vein location. On the remaining 3 patients, the effective anticoagulation was contra-indicated because of a haemorrhagic cerebral transformation (n = 2) or by a digestive bleeding (n = 1), leading to the use of a cava filter in one patient and of thromboprophylaxis for all. Elastic stocks were used in all cases to prevent DVT.

## Discussion

The main finding of our study is a high prevalence of 38.5% of asymptomatic DVT (n = 5) in patients with neurovascular diseases and non-severe COVID-19 in our stroke unit. To our knowledge this is the first study exploring venous lower limbs with systematic bedside DUS. More importantly, this rate is as important in our study as in patients with severe COVID-19 and admitted to intensive unit care [3, 4]. This non-invasive procedure may be more accurate to detect DVT than D-dimer levels, which is correlated to COVID-19 severity [7], because they are also increased in the acute phase of stroke with no COVID-19 [8]. Our findings show rates that are more than three times those reported in the stroke literature [8] and suggest that non-severe COVID-19 may increase risk for DVT.

Among the factors increasing the risk of DVT in stroke patients it was shown that taken together a higher age, a higher NIHSS before and after treatment, and a longer hospitalization are deleterious [9, 10]. In one hand our findings confirm these results but in the other hand they show a discrepancy since in acute stroke these authors found only 11.5% DVT patients [9]. Literature in severe COVID-19 reports a systemic inflammation induced by a cytokine storm favouring a state of hypercoagulability [11,12], which in turn may be a risk factor of DVT, but it seems overestimated [13]. It was also reported however that the viral infection in COVID-19 affects the venous endothelial cells [14] and induces an endotheliitis [15], which in turn leads to endothelial dysfunction predisposing to pro-coagulant state [16]. Therefore, we hypothesized that in non-severe COVID-19 patients with a neurovascular disease treated with thromboprophylaxis, the peripheral venous thrombotic complications may appear more frequently than in non-COVID patients. Further studies are needed to confirm this hypothesis by comparing data from COVID-19 patients with those of non-COVID-19 patients.

## Conclusion

We showed for the first time an important prevalence of 38.5% DVT using systematic bedside DUS in neurovascular disease patients with non-severe COVID-19 despite recommended thromboprophylaxis and justified by the absence of reliable biological parameters. We hypothesize that COVID-19 increase the risk of DVT in stroke patients. Since it was suggested that DUS should be performed in all patients and integrated into medical care [3, 4] of COVID-19 patients and considering the risk of DVT in stroke patients and COVID-19 as an additional risk factor for thrombosis [12] we strongly suggest a routine DUS of lower limbs in such situation.

## Data Availability

the data mentioned in the manuscript are available on request to the corresponding author.

## Acknowledgement

The authors would thank R Galani PhD for his writing assistance.

## Author Contributions

OR designed the study and wrote the manuscript. PPIN, BAT, JD, and BD obtained data. BG and WV critically reviewed the manuscript.

## Table Legend

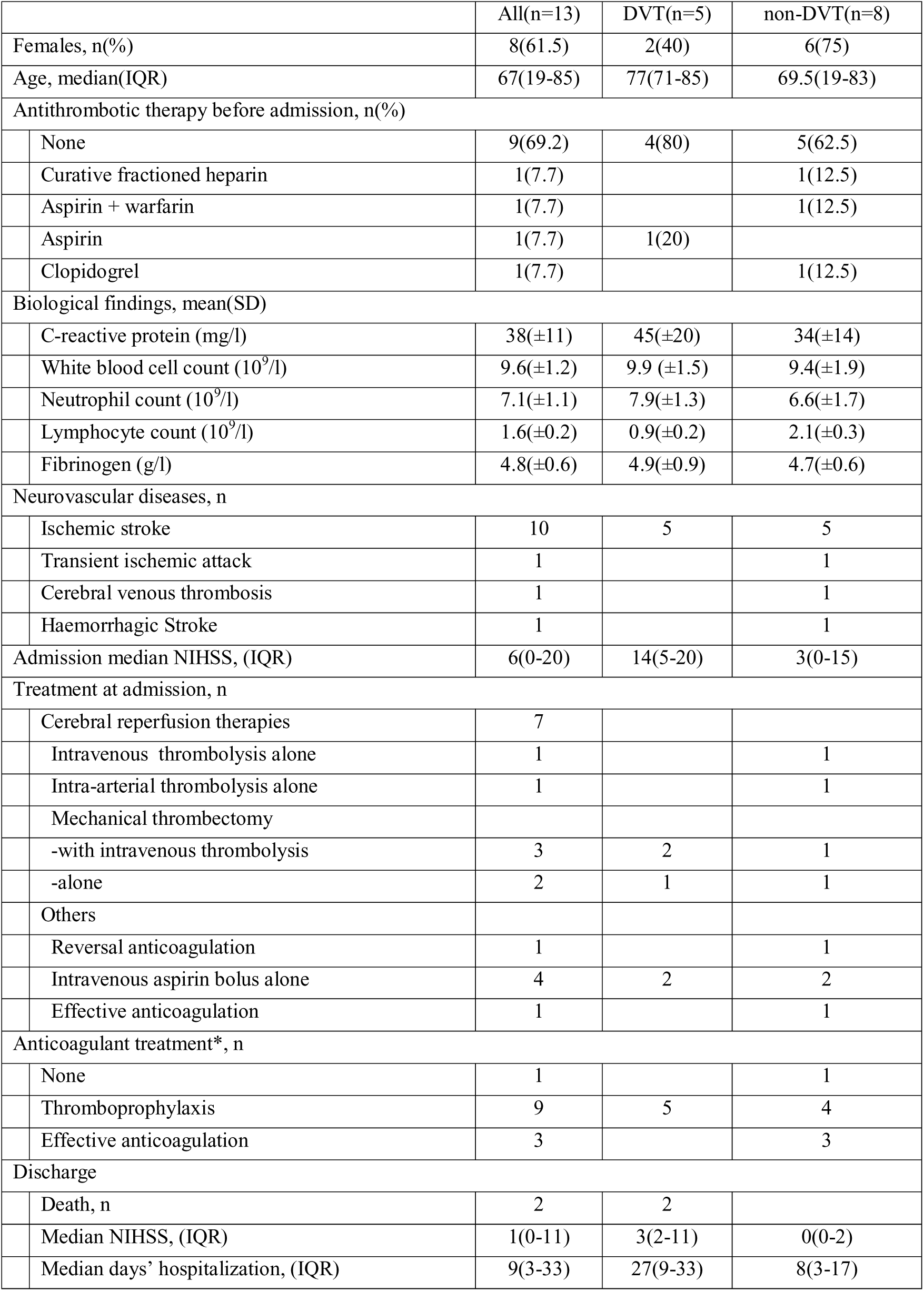
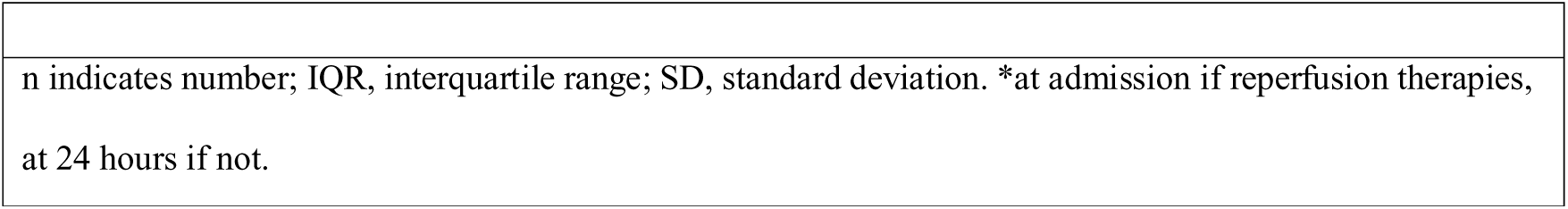
Characteristics of neurovascular diseases patients with and without DVT and COVID-19.

|  | All(n=13) | DVT(n=5) | non-DVT(n=8) |
| --- | --- | --- | --- |
| Females, n(%) | 8(61.5) | 2(40) | 6(75) |
| Age, median(IQR) | 67(19-85) | 77(71-85) | 69.5(19-83) |
| Antithrombotic therapy before admission, n(%) |  |  |  |
| None | 9(69.2) | 4(80) | 5(62.5) |
| Curative fractioned heparin | 1(7.7) |  | 1(12.5) |
| Aspirin + warfarin | 1(7.7) |  | 1(12.5) |
| Aspirin | 1(7.7) | 1(20) |  |
| Clopidogrel | 1(7.7) |  | 1(12.5) |
| Biological findings, mean(SD) |  |  |  |
| C-reactive protein (mg/l) | 38(±11) | 45(±20) | 34(±14) |
| White blood cell count (10<sup>9</sup>/l) | 9.6(±1.2) | 9.9 (±1.5) | 9.4(±1.9) |
| Neutrophil count (10<sup>9</sup>/l) | 7.1(±1.1) | 7.9(±1.3) | 6.6(±1.7) |
| Lymphocyte count (10<sup>9</sup>/l) | 1.6(±0.2) | 0.9(±0.2) | 2.1(±0.3) |
| Fibrinogen (g/l) | 4.8(±0.6) | 4.9(±0.9) | 4.7(±0.6) |
| Neurovascular diseases, n |  |  |  |
| Ischemic stroke | 10 | 5 | 5 |
| Transient ischemic attack | 1 |  | 1 |
| Cerebral venous thrombosis | 1 |  | 1 |
| Haemorrhagic Stroke | 1 |  | 1 |
| Admission median NIHSS, (IQR) | 6(0-20) | 14(5-20) | 3(0-15) |
| Treatment at admission, n |  |  |  |
| Cerebral reperfusion therapies | 7 |  |  |
| Intravenous thrombolysis alone | 1 |  | 1 |
| Intra-arterial thrombolysis alone | 1 |  | 1 |
| Mechanical thrombectomy |  |  |  |
| -with intravenous thrombolysis | 3 | 2 | 1 |
| -alone | 2 | 1 | 1 |
| Others |  |  |  |
| Reversal anticoagulation | 1 |  | 1 |
| Intravenous aspirin bolus alone | 4 | 2 | 2 |
| Effective anticoagulation | 1 |  | 1 |
| Anticoagulant treatment*, n |  |  |  |
| None | 1 |  | 1 |
| Thromboprophylaxis | 9 | 5 | 4 |
| Effective anticoagulation | 3 |  | 3 |
| Discharge |  |  |  |
| Death, n | 2 | 2 |  |
| Median NIHSS, (IQR) | 1(0-11) | 3(2-11) | 0(0-2) |
| Median days' hospitalization, (IQR) | 9(3-33) | 27(9-33) | 8(3-17) |

|  |
| --- |
| n indicates number; IQR, interquartile range; SD, standard deviation. *at admission if reperfusion therapies, at 24 hours if not. |

